# Seroprevalence and Risk Factors of Chikungunya in Ethiopia: A Systematic Review and Meta-Analysis

**DOI:** 10.1101/2024.01.04.24300817

**Authors:** Gashaw Getaneh Dagnaw, Abebe Tesfaye Gessese, Mebrie Zemene Kinde, Abebe Belete Bitew, Haileyesus Dejene, Bereket Desalegn, Solomon Lulie Abey

## Abstract

The recurrence of Chikungunya virus poses a significant public health concern, given its association with numerous epidemic episodes in Africa, Asia, and India. Since the virus was first detected in Ethiopia in 2016, the disease has been identified in different parts of the country, emphasizing the need for up-to-date epidemiological data on the status of Chikungunya in Ethiopia. We conducted a systematic review and meta-analysis using online published articles between 2016 and 2023 from PubMed, Scopus, and Google Scholar databases. The PRISMA guideline was thoroughly followed and registered in the Prospective Register of Systematic Reviews (PROSPERO). A set of keywords like ‘Chikungunya,’ ‘Chikungunya Virus,’ ‘Chikungunya Fever,’ ‘Prevalence,’ ‘Seroprevalence,’ “Risk Factor”, “Potential Factors”, and ‘Ethiopia’ were used in the search engines. A total of five articles met the eligibility criteria and were included for data extraction. Meta-analysis was performed using STATA 17 software. The pooled seroprevalence of Chikungunya in Ethiopia was 12.35%. The highest prevalence was reported in Southern Nations, Nationalities, and Peoples’ Region (SNNPR) at 43.6%, while the lowest seroprevalence was in Dire Dawa, at approximately 12%. Factors such as occupation, education, age, and sex contributed to prevalence variation. Subgroup meta-analysis showed heterogeneity across the types of studies employed. No indications of publication bias or small-study effects were found in the Begg’s test and Egger’s test. The findings will help us to understand the trend of Chikungunya in Ethiopia. The findings recommend proactive monitoring or active surveillance of viral diseases and the rigorous enforcement of health systems, as well as One Health measures in Ethiopia, to improve human public health.

## INTRODUCTION

Chikungunya, a mosquito-borne viral disease, is caused by an RNA virus belonging to the Alphavirus genus of the family Togaviridae, known as Chikungunya virus (CHIKV). It is responsible for millions of documented cases worldwide. While the majority of cases recover within weeks, joint and musculoskeletal pain can persist for months to years’ post-infection (1). The disease is characterized by clinical signs such as fever, debilitating severe joint pain, joint swelling, muscle pain, headache, nausea, fatigue and rash (2).

Chikungunya virus was first isolated during a 1952–53 outbreak in southern Tanzania (3), although the clinical descriptions suggesting its presence as far back as the 1600s (4). Today, CHIKV has become widespread globally, identified in more than 110 countries and represents a significant global public health concern (5). The virus has expanded its geographic range, causing outbreaks in various regions around the world and posing a global public health concern. Factors such as climate change, vector adaptations, urbanization, and human migration have contributed to the spread of the virus to new areas, including regions beyond its historically endemic zones (6).

In addition, studies showed that Chikungunya prevalence varies with factors such as occupation, age, sex, and education. Farmers and aged populations are associated with higher prevalences. Gender disparities also play a role, influencing exposure and transmission patterns (7,8).

Chikungunya in Ethiopia is becoming a significant public health concern, as it has caused considerable morbidity since first being detected (9). The virus was first documented in Ethiopia in June 2016, with the confirmation of its first case in the Suuf kebele, Dollo Ado district of the Somalia regional state (10) that assumed to be originated from Kenya. Somalia regional state shares a border with the Mandera region of Kenya, where a Chikungunya outbreak was ongoing (11). Since then, Chikungunya has spread rapidly and has been reported in different districts of Ethiopia (12,13).

At present, there is no approved vaccine or antiviral therapies available for Chikungunya (14). However, the approach for the treatment, and control and prevention of the diseases is through alleviating the symptoms or supportive treatment, and eliminating the mosquitoes, that transmit the virus (14,15). In addition, public awareness creation through community education and training about the outbreak of emerging and reemerging vector-borne diseases, methods of transmission, and their control and prevention methods remain an important mechanism for managing Chikungunya (14).

Rapid and accurate diagnosis is the prerequisite approach for implementation of control and prevention of the disease outbreak although there are limitations in laboratory and public health infrastructure (16). Ethiopia, like many developing nations, struggles with a range of public health challenges that contribute to the outbreak of diseases. Limited healthcare infrastructure and uneven distribution of resources hinder effective prevention, detection, and response to health crises (16).

Despite the limitations, Ethiopia made an effort to limit the spread of CHIKV across the country and to prevent the potential transmission of the disease in the affected regions (9). The government was taking vector control measures such as indoor residual spraying, distributing insecticide-treated bed nets and encouraging the population to eliminate a breeding ground for mosquitoes like stagnant water (17).

However, the effectiveness of these measures may vary, and there are reports of the disease from various locations of Ethiopia. The disease’s current status in Ethiopia is unclear. Knowing the up-to-date epidemiological data of the disease is important for preparedness and implementation of control and prevention of the Chikungunya in Ethiopia. Therefore, this systematic review and meta-analysis aimed to provide current information on Chikungunya diseases and providing valuable insights for health professionals and concerned authorities to prepare effective control and prevention strategies.

## METHODS

### Systematic Review Protocols

The guidelines and procedures of the Preferred Reporting Items for Systematic Reviews and Meta-Analysis (PRISMA) (18), were followed in this systematic review and Meta-analysis (Figure 1), and registered in the database of Prospective Register of Systematic Reviews (PROSPERO) under the reference number CRD42023271579.

**Figure 1.**
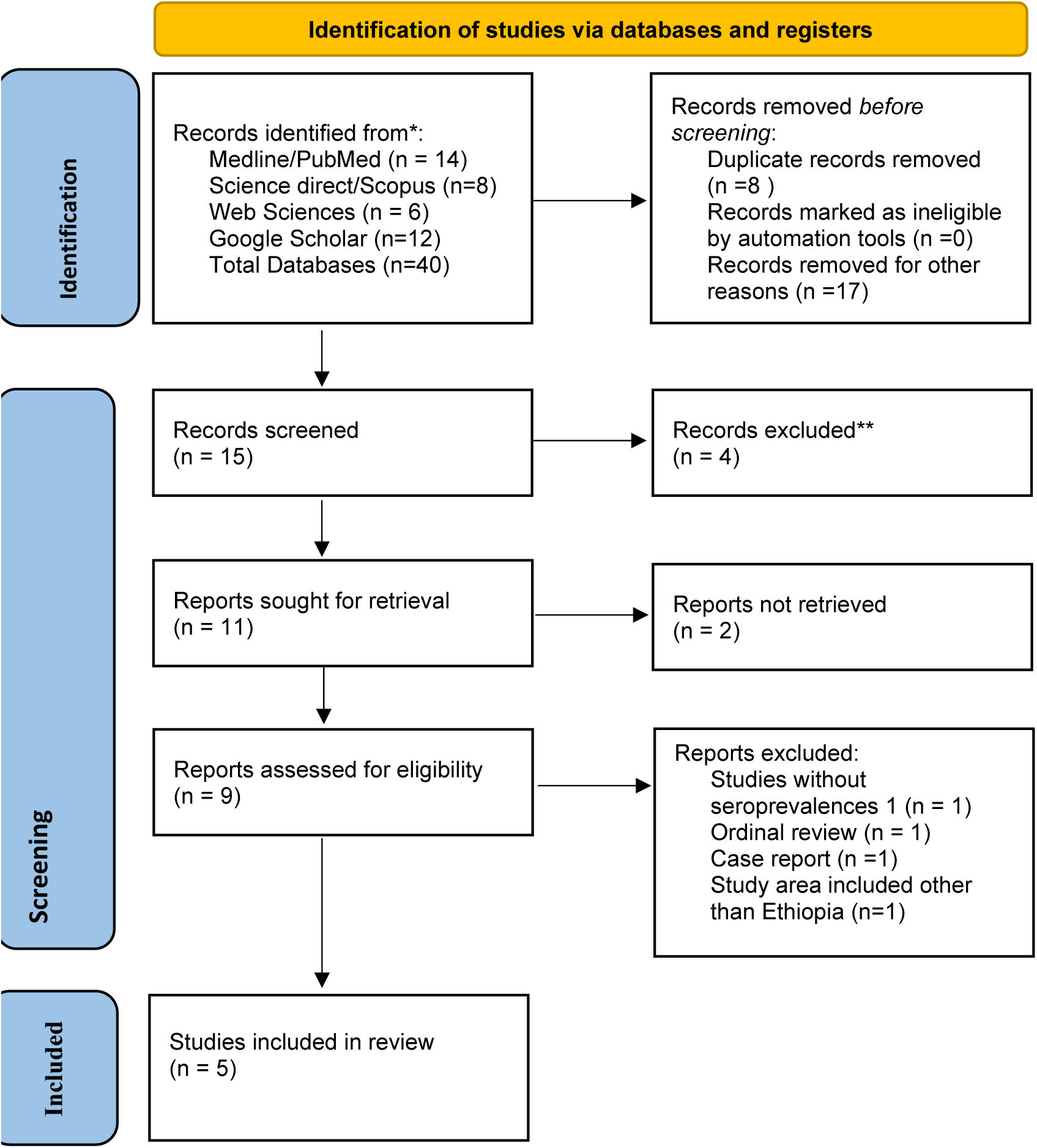
Preferred Reporting Items for Systematic Reviews and Meta-Analysis (PRISMA) 2020 flow diagram presents the search strategy and article selection process published between 2016 and 2023, for the current study. Based on the search criteria, a total 40 publications were identified and through the refinement steps outlined in the PRISMA flowchart, the final selection resulted in five full-text articles in the English language that were included in this systematic review.

### Literature Search and Sources

The data were extracted from published public articles available from different electronic databases including Medline/PubMed, Science Direct/Scopus, Google scholar and Web of Science. A sets of keywords like “Chikungunya”, “Chikungunya Virus”, “Chikungunya Fever”, “Vector-borne”, Arbovirus, “Incidence”, “Prevalence”, “Seroprevalence”, “Seroepidemiology”, “Risk Factors”, “Potential Factors” and” Ethiopia” were used on the search. The search queries were configured using Medical Subject Headings (MeSH) and the “OR and “AND” Boolean operators were used to identified studies with any of the keywords in their titles, abstracts and full texts that might be included on this systematic and meta-analysis review. Moreover, unpublished thesis manuscripts were also accessed from various Ethiopian Universities and research centers.

### Eligibility Criteria

Observational studies focused on CHIKV infection or disease conducted in Ethiopia and involving the general population or specific age group were included in this study. Original articles published online in the English language between 2016 and November 2023 were included to ensure the incorporation of updated information on the issue. The eligible studies were those which provided the seroprevalence of Chikungunya in case-control, cross-sectional and cohort studies that reported lab-confirmed cases relied on laboratory tests, encompassing serologic or virological antibodies detection methods such as ELISA IgG, ELISA IgG + IgM and/ or molecular diagnosis. The title and abstracts of articles were initially examined to ascertain whether they provided essential information, including the total sample size and the reported outcome of interest (number of positive samples). Subsequently, the full text was assessed. Duplicates and those that did not contain all relevant information for this systematic review and meta-analysis were removed. Case report studies and reviews were utilized exclusively as reference sources and were not included in the comprehensive analysis of this systematic review. The analysis did not take into account the specificity and sensitivity of the tests.

### Study Selection and Data Extraction

Two experts (ATG and SLA) independently identified articles from the search engines using key terminologies and subsequently screened them based on their titles and abstracts. Selected publications were then imported into Mendeley, and their full texts were retrieved. Eligibility of the full texts was assessed by checking if they addressed the main outcomes of interest. The proportion of the Seropositive of Chikungunya was considered as the main outcome of the study. Data extraction took place independently by two experts (GGD and MZK) from November 1, 2023, to November 21, 2023, and cross-verification was performed by another two experts (HD & BD). If discrepancies arose, the data were re-extracted, even though there were no initial discrepancies.

### Quality and Risk-of-Bias Assessment

The quality of the selected articles was evaluated based on PRISMA quality assessment checklist (PRISMA 2020 Checklist) (18), which included items assessing objectives, different components of the methods (e.g., study design, sample size, study population, bias, statistical methods), results, and limitations (S1 Table). This quality assessment was done by two experts (ABB & GGD). In addition, the presence of publication bias or small-study effects was assessed by using Begg’s test and Regression-based Egger test, which examines the correlation between the effect size and the standard error of the effect size across studies.

### Meta-analysis and Data Analysis

The extracted data was entered into an Excel spreadsheet 2019. The seroprevalence of Chikungunya from each study was recorded and the individual study weight, standard error, and 95% confidence interval (CI) were calculated based on the inverse variance method and the binomial equation (19). The logit transformation of proportional prevalence with its variance and standard error was calculated. Subgroup analyses for the primary outcome (seroprevalence of Chikungunya) was done using DerSimonian and Laird model by considering geographical locations, and laboratory techniques employed (PCR or ELISA) (20). An estimate of heterogeneity among and within the studies was calculated from the inverse variance of the random effects model (21). The parameter tau-squared (τ^2^), I-squared (I^2^) and H-squared (H^2^) were calculated to measure the variance of the true effect sizes between-study variance, inter-study heterogeneity, and total variability, respectively (22). Pooled prevalence and standard error with 95% confident interval (CI) were calculated.

Reports containing seroprevalence data were summarized in a table, organized by reference and study area. This table includes details on study design, study area, sample size, and test type. Descriptive statistics were employed for data summarization, and the findings were illustrated through figures and tables. STATA version 17 software was employed to perform the statical tasks.

## RESULTS

### Study Characteristics and Search Results

Figure 1 depicts the complete procedures illustrating how the included articles were selected. A total of 40 articles involving around 335821 subjects published between 2016 and 2023 were identified using search engines. Initially, 17 of them were excluded during identification of articles by titles for their lack of relevance to the review. The combination of reference manager software (Mendeley) and manual confirmation was employed to identify and eliminate 8 duplicated papers. Subsequently, 15 articles underwent screening but only 9 articles underwent evaluation according to eligibility criteria. Only five publications met the eligibility criteria and were included in the final systematic review and meta-analysis (Figure 1). The selected articles encompassed research conducted in five regional states: South Nations and Nationalities Region, Amhara, Tigray, Gambella, and South Eastern Ethiopia, as well as one city administration (Dire Dawa) (Figure 2). The included studies comprised cross-sectional (80%, 4/5) and one case-control study (20%, 1/5) (Table 1).

**Figure 2.**
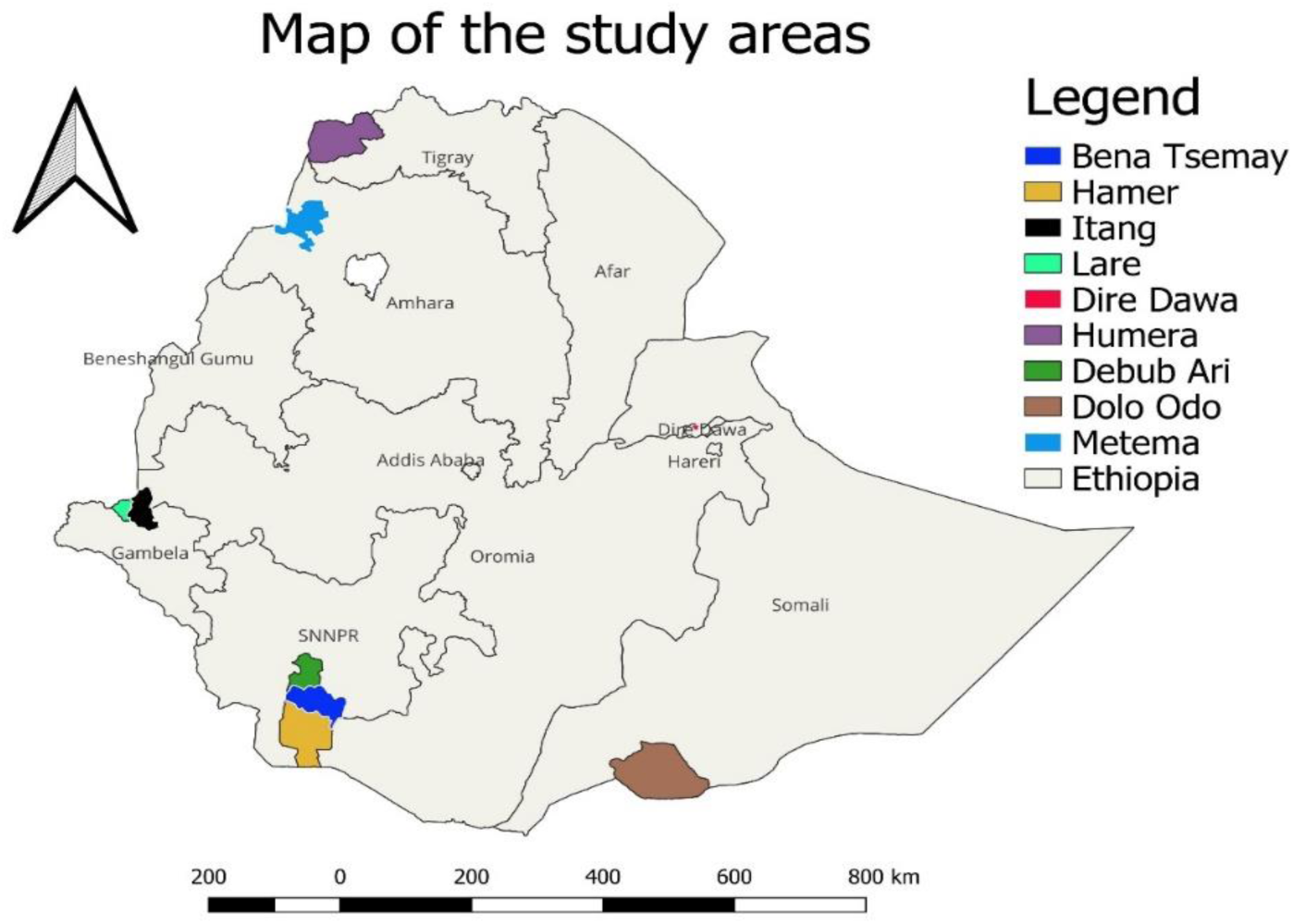
Map of Ethiopia showing Chikungunya virus disease in selected publications between 2016 and 2023 (QGIS version 3.34.1 was used for the map).

**Figure 3.**
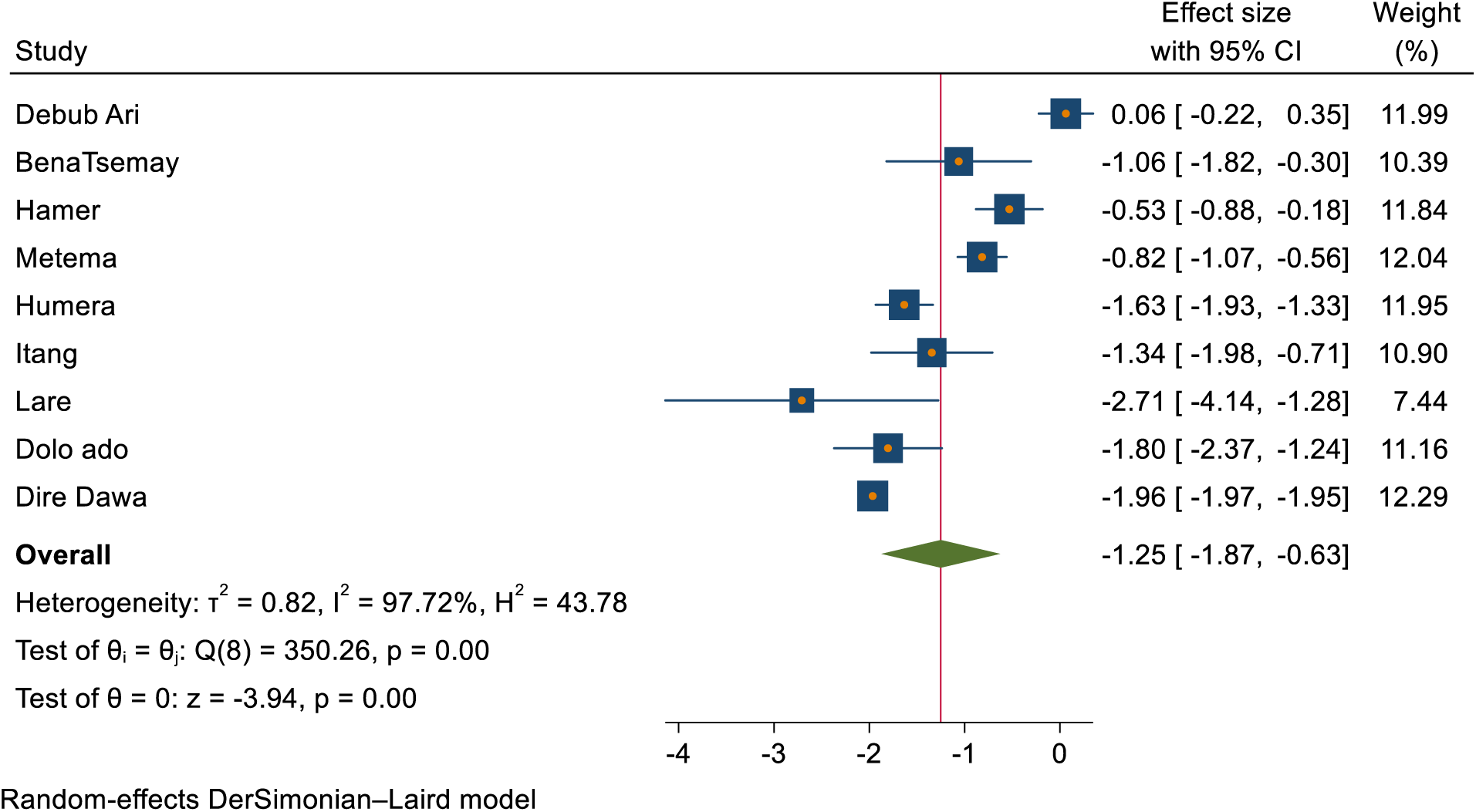
The random effect estimates of the effect size in different districts of Ethiopia.

**Figure 4.**
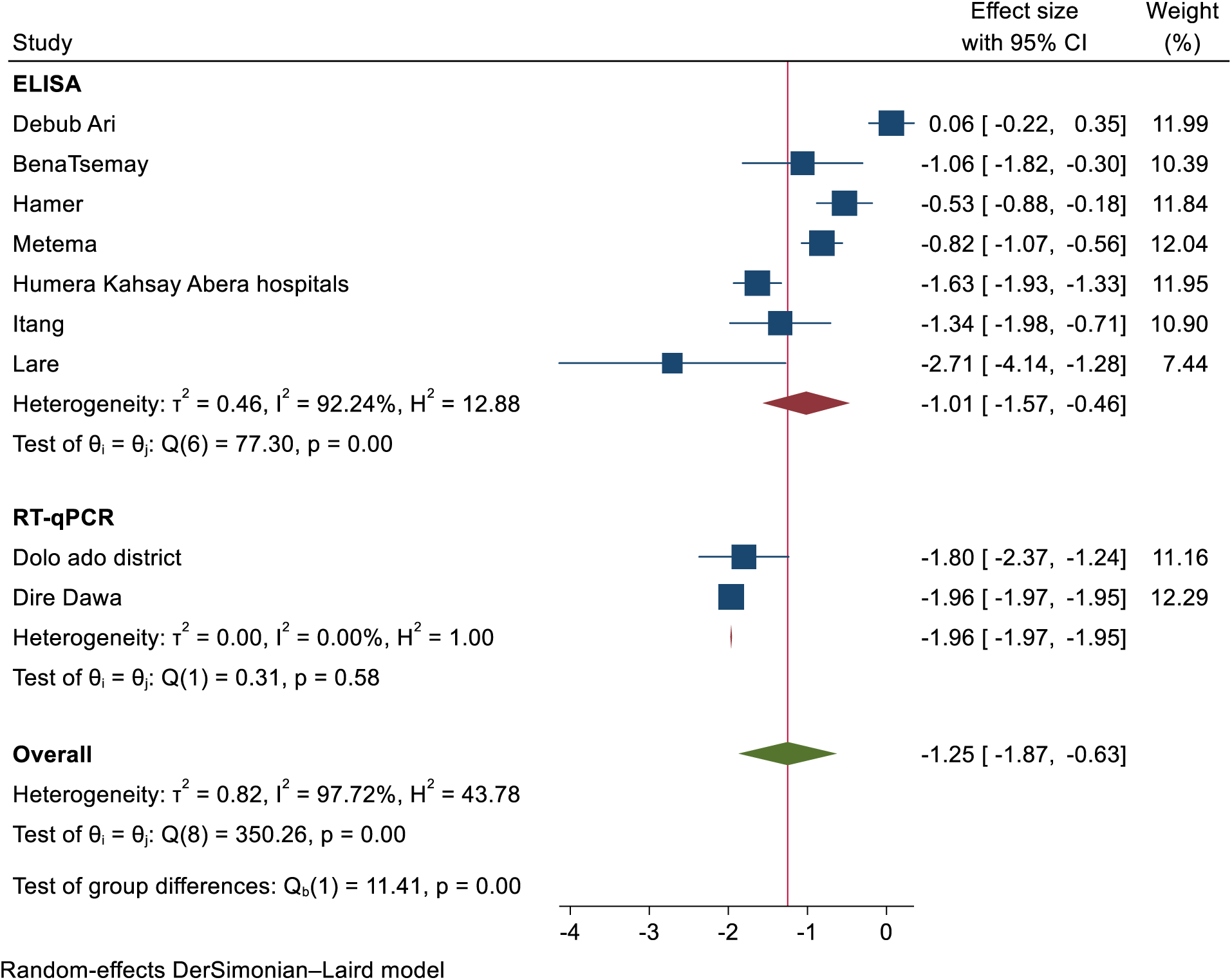
The random effect estimates of the effect size across the type of test performed.

**Table 1:** The seroprevalence of Chikungunya disease in Ethiopia from selected publications between 2016 and 20120.

| Authors and study year | Study area |  | Study design | Sample size | Test type | Sero-prevalences (%) | logit (95% CI) |
| --- | --- | --- | --- | --- | --- | --- | --- |
|  | Region | District |  |  |  |  |  |
| Endale et al., 2020 | SNNPR | Debub Ari | Cross-sectional | 190 | ELISA | 51.58 | 0.15 (-0.22, 0.35) |
|  | SNNPR | BenaTsemany |  | 35 | ELISA | 25.71 | 0.39 (-1.82, -0.30) |
|  | SNNPR | Hamer |  | 135 | ELISA | 37.04 | 0.18 (-0.88, -0.18) |
| Ferede et al., 2021 | Amhara | Metema | Cross-sectional | 274 | ELISA | 30.66 | 0.13 (-1.07, -0.56) |
|  | Tigray | Humera |  | 312 | ELISA | 16.35 | 0.15 (-1.93, -1.33) |
| Asebe et al., 2021 | Gambella | Itang | Cross-sectional | 58 | ELISA | 20.69 | 0.32 (-1.98, -0.71) |
|  | Gambella | Lare |  | 32 | ELISA | 6.25 | 0.73 (-4.14, -1.28) |
| Takele, 2020 | South-Eastern Ethiopia | Dolo ado | Case-control | 99 | RT-qPCR | 14.14 | 0.29 (-2.37, -1.24) |
| Geleta et al., 2019 | Dire Dawa | Dire Dawa | Cross-sectional | 334686 | RT-qPCR | 12.30 | 0.01 (-1.97, -1.95) |

CI = Confidence Interval

There were two types of assays used for diagnosis across the studies namely, Enzyme-Linked Immunosorbent Assay (ELISA; 80%, 4/5), and Quantitative reverse transcription polymerase chain reaction (RT-qPCR; 20%,1/5) (Table 1).

### Seroprevalence of Chikungunya

The pooled seroprevalence of Chikungunya in Ethiopia was 12.35% (95%CI: –0.57, 0.81). The highest prevalence was reported in the Southern Nations, Nationalities, and Peoples’ Region (SNNPR) at 43.6%, while the lowest seroprevalence was found in Dire Dawa, with approximately 12%. At district level, the highest Chikungunya infection occurred in the Bebub Ari district at 51.58% (SE (logit)=0.15, 95%CI=-0.22, 0.35) while the lowest prevalence was recorded from the Lare district at 6.25% (SE (logit)=0.73, 95%CI=-4.14, –1.28) (Table 1). Look S1 figure 2 for the regions and districts of the study area.

### Risk Factors Associated with Chikungunya Infection

Risk factors, include occupation, education, sex, and age were identified in four of the included articles. The Chikungunya prevalence exhibited variation based on these factors. Notably, the prevalence of Chikungunya was significantly varied with occupation, irrespective of geographical location. According to Endale et al. (2020), farmers showed the highest seroprevalence of Chikungunya at 49.7%, compared to pastoralists at 34.9%. Similar findings were reported in northwest Ethiopia by Ferede et al. in 2021. Additionally, Asebe et al. (2021) reported that pastoralists had the lowest Chikungunya infection rate at 4.1% (see Figure 5).

**Figure 5.**
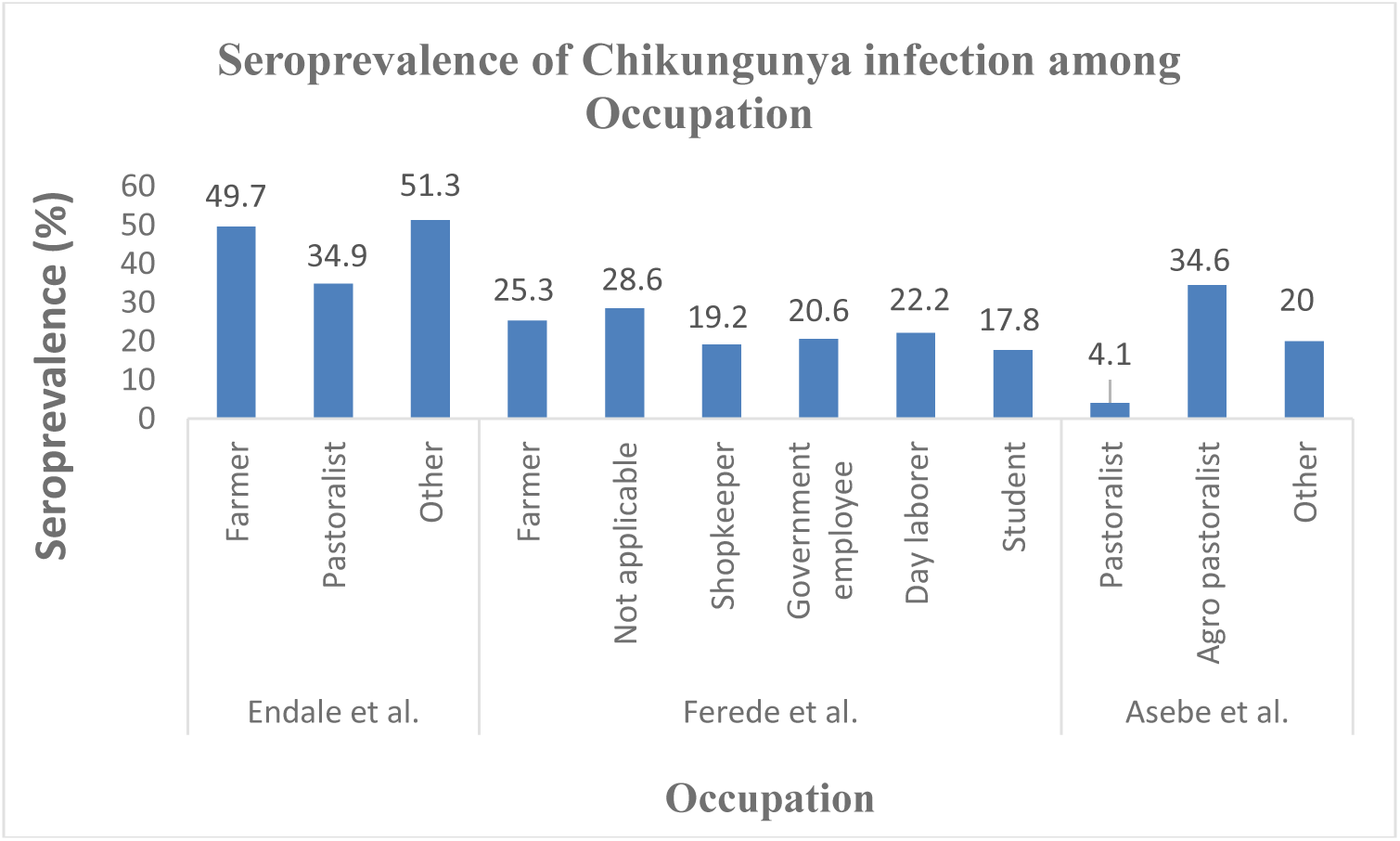
Distribution of Chikungunya disease among farmers, pastoralists, agro-pastoralists, and individuals in other occupations in selected publications from 2016 to 2023.

Figure 6 displays the prevalence of Chikungunya across different educational backgrounds. Chikungunya prevalence was higher among individuals who had received formal education compared to those who had not attended formal education or were illiterate (see Figure 6).

**Figure 6.**
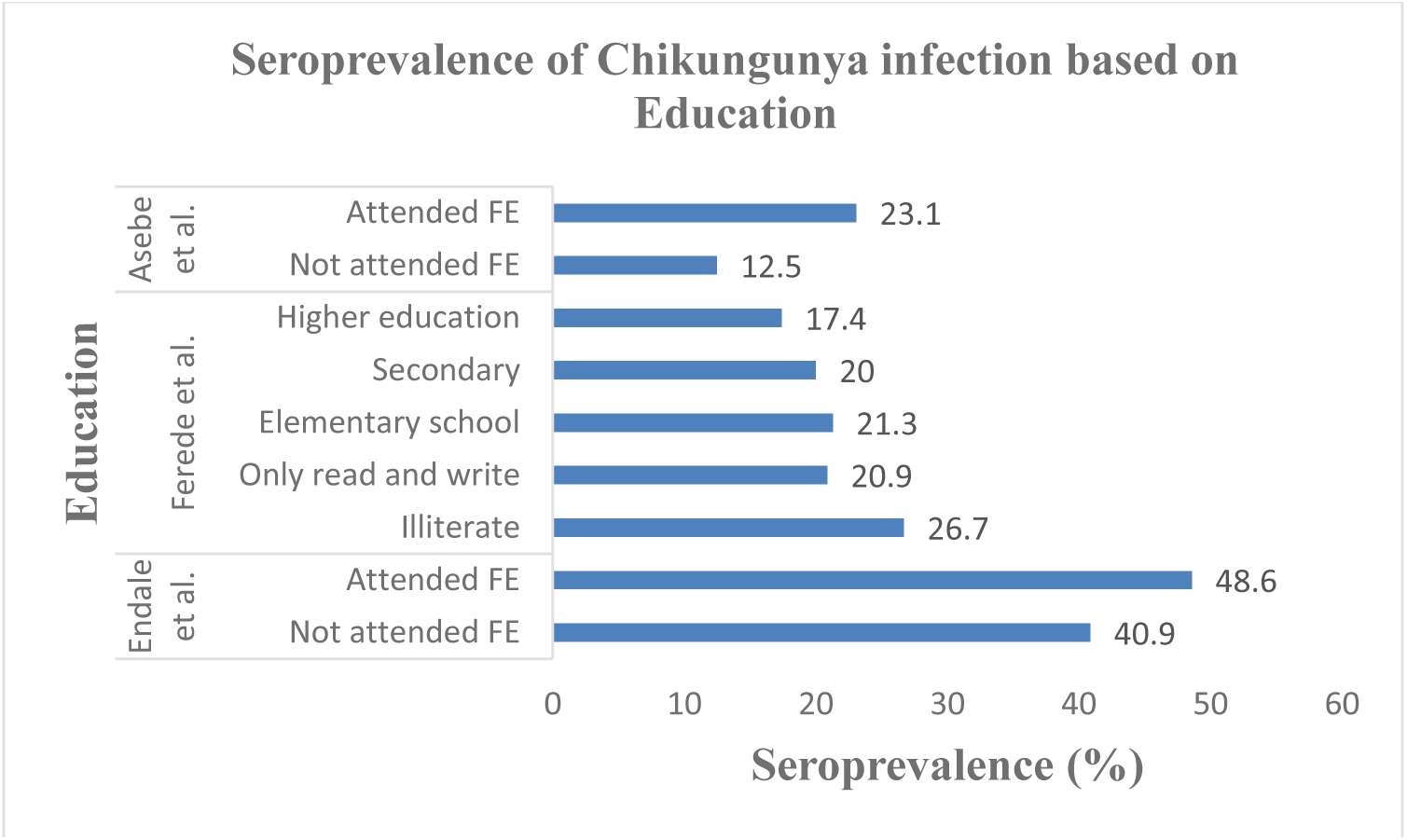
Distribution of Chikungunya disease in people’s educational status: attended formal education (FE), Not attended FE, higher education, secondary, elementary school, only read and write, and illiterate in selected publications between 2016 and 2023.

The seroprevalence of Chikungunya was higher in the adult age group, while the lowest prevalence was recorded in children. Endale et al. (2020) reported that the highest Chikungunya prevalence in the age group of 36-55 years at 53.5% and the lowest was approximately 17.7% in the age group of 5-10 years. Similar results were reported by Ferede et al. (2021). However, Geleta et al. (2019) reported a higher Chikungunya infection rate in the age group of 5-14 years at 17.1%, with the lowest prevalence in the age group of ≤5 years at 3.6% (see Figure 7).

**Figure 7.**
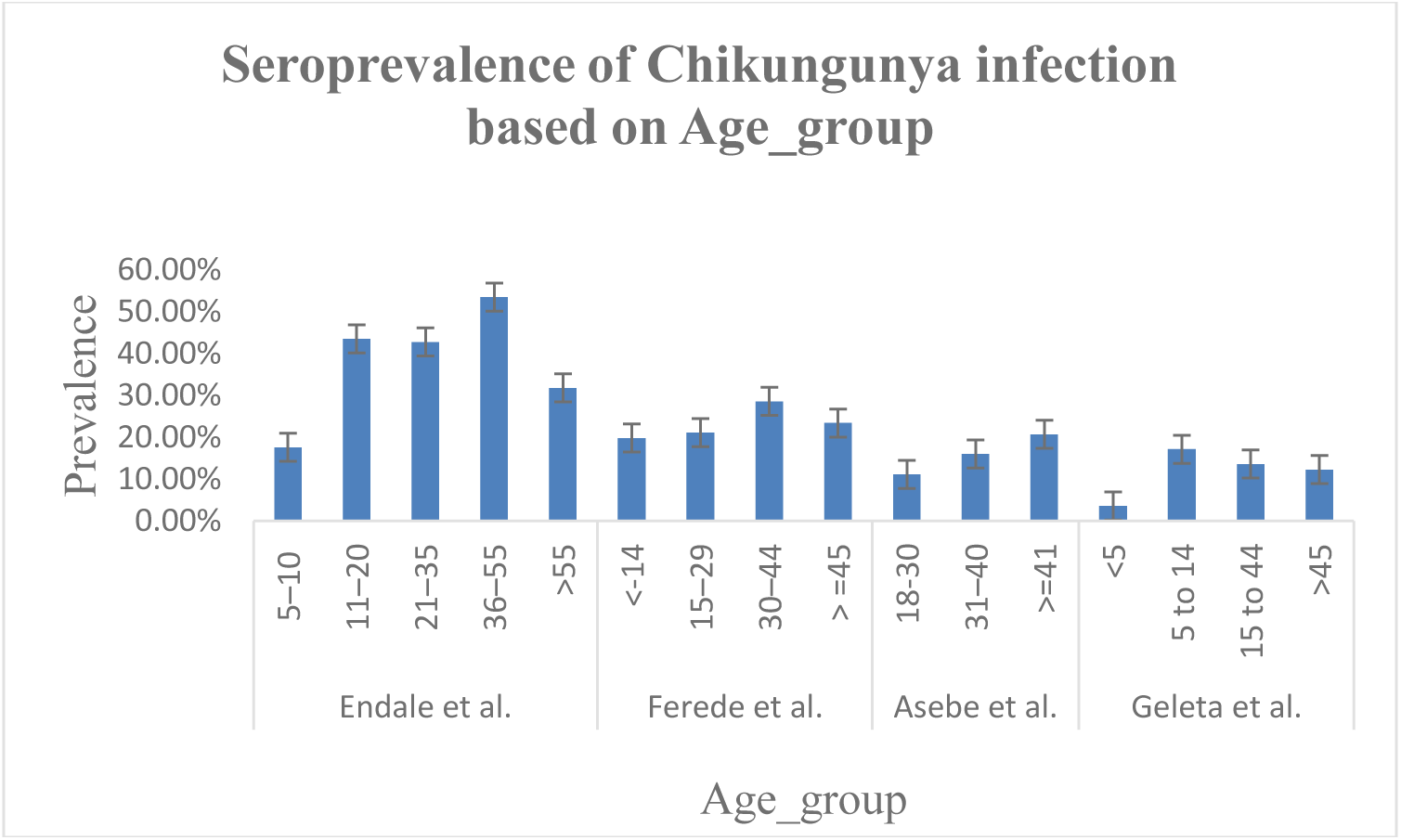
Distribution of Chikungunya disease in different age group of the participants in selected publications from 2016 to 2023.

Figure 8 showed the prevalence of Chikungunya in males and females. It is notable that the seroprevalence of Chikungunya across different sex groups was not consistent. Asebe et al. (2021) reported a higher Chikungunya infection rate in males at 22.64%, compared to females at 5.41%. Similarly, Ferede et al. (2021) found a higher prevalence in males, with 26.8%, compared to 14.14% in females. However, Endale et al. 2020) reported a higher prevalence among females at 47.7% compared to males at 39.78%. While approximate seroprevalence of Chikungunya in both sexes was reported by Geleta et al. (2019) (see Figure 8).

**Figure 8.**
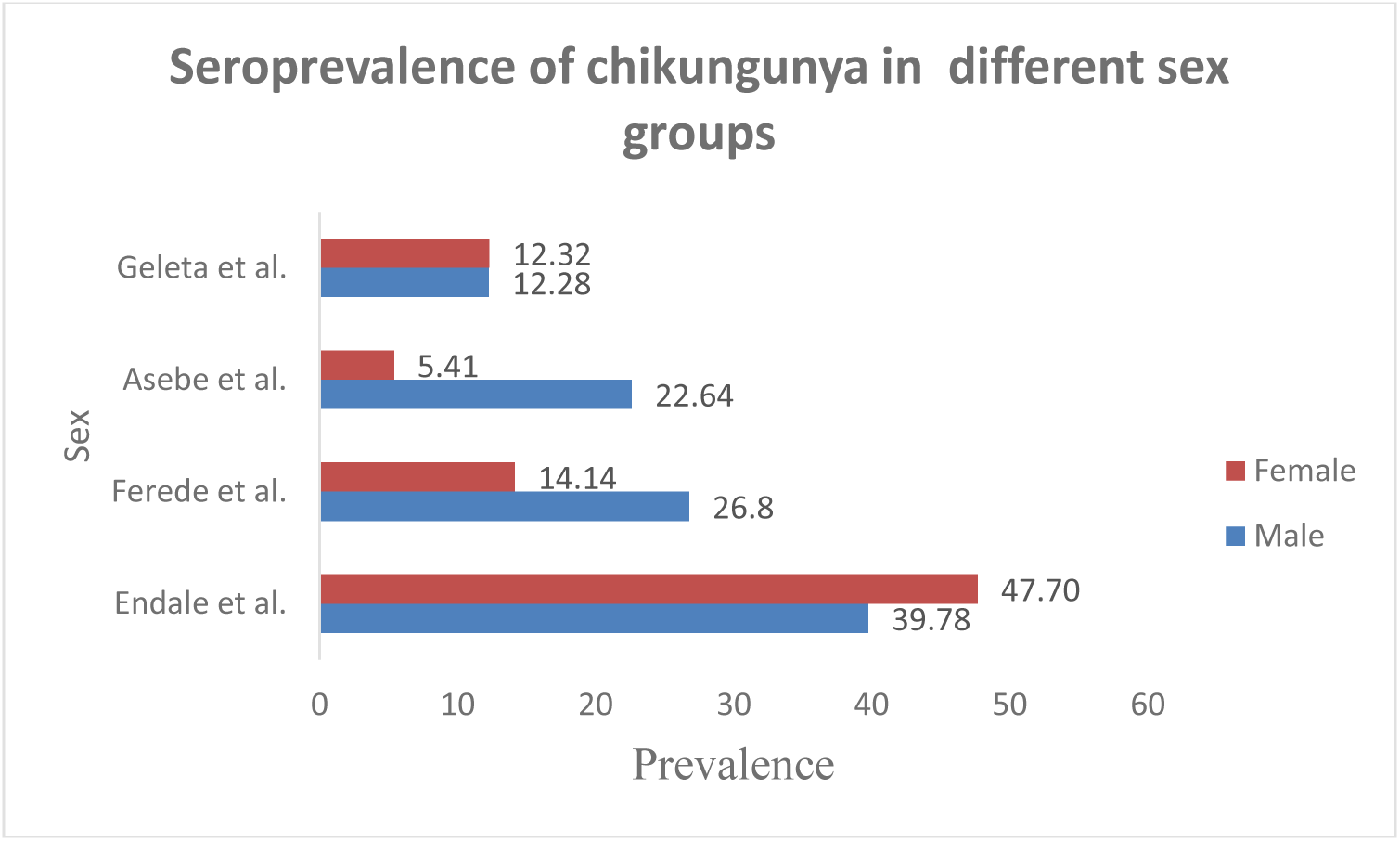
The prevalence of Chikungunya disease in Ethiopia in males and females in selected publications between 2016 and 2023.

### Meta-analysis

A forest plot generated by the DerSimonian–Laird model using a Random Effects approach at the district level resulted in negative effect size. Districts such as Dire Dawa and Metema exhibited considerable negative effect size but had higher weight (Effect size (ES)=-1.96, 95%CI –1.97, –1.95, weight (W)=12.29%) and (ES=-0.82, 95%CI=-1.07, – 0.56, W=12.04), respectively. While Lare and Bena Tsemay had considerable negative effect size but contributed the lowest (–2.71, 95%CI –4.14, –1.28, W=7.44%), and (ES=-1.06, 95%CI=-1.82, –0.30, W=10.39%), respectively. Debub Ari had positive effect size of 0.06 (95%CI=-0.22,0.35, W=11.99%). The overall random effect estimates indicate a significant negative effect size of –1.25 (95% CI= –1.87, –0.63), indicated an overall lower seroprevalence of Chikungunya across the districts (Figure 3). There was statically significant variability among the districts (τ^2^=0.82, I^2^= 97.72%, H^2^ = 43.78, p=0.00).

The subgroup analysis based on the type of tests conducted revealed variations in the study outcomes. The test Enzyme-Linked Immunosorbent Assay (ELISA) was showed the variability among the studies and the heterogeneity was statically significant (I^2^ = 92.24%, p=0.00). Whereas the test Reverse Transcription quantitative Polymerase Chain Reaction (RT_qPCR) revealed lack of variability among the studies (I^2^ = 0.00%, p= 0.58). The pooled effects size estimate –1.25 (–1.87, –0.63) indicated that there was a heterogeneity between ELISA and RT-qPCR tests with I^2^ = 97.72% (p = 0.00) (see Figure 4).

### Quality assessment

The statistical tests for small-study effects in a meta-analysis resulted in non-significant results. The Begg’s test showed the absence of significant small-study effects (Kendall’s Score = –8.00, SE of Score=9.592, z-value=-0.94, P=0.47). similarly, the regression-based Egger test detected funnel plot asymmetry and revealed no indication of publication bias or small-study effects in the meta-analysis (beta1= –1.74, SE of beta1= 1.766, z-value= –0.99, p=0.32). Look S1 Figure, Galbraith plot also showed the absence of small-study effect.

## DISCUSSION

Five studies on Chikungunya conducted in Ethiopia and published in English between 2016 and 2023 were deemed eligible and included in this systematic review and meta-analysis. The meta-analysis resulted in a pooled seroprevalence of Chikungunya at 12.35%. Factors such as geographical location, occupation, age, sex, and education contributed to the variation in Chikungunya seroprevalence. Subgroup analysis based on the study area and the type of tests performed revealed significant heterogeneity.

The pooled prevalence of Chikungunya highlights the significance of the disease in Ethiopia. This prevalence was relatively lower than reports from neighboring countries. For instance, Sudan showed a seroprevalence among general populations of median 12%, with a range of 0-43% (8), while Kenya exhibited prevalence rates ranging from 0.97% to 42% (11). This disparity prompts an exploration of potential contributing factors. Geographical nuances, including climate and ecosystems, may impact the abundance and behavior of the Aedes mosquitoes, which are the vectors responsible for CHIKV transmission (17). Furthermore, variations in sanitation and hygiene practices, as well as the effectiveness of vector control measures, could influence the prevalence of Chikungunya. Another aspect to consider is the diagnostic capacity; differences in the accuracy and sensitivity of disease detection methods could lead to variations in reported prevalence.

In this systematic review and meta-analysis, the highest seroprevalence of Chikungunya was reported in the Debub Ari district of SNNPR, while the smallest was recorded in the Lare district of the Gambella regional state. In addition, seroprevalences differed among districts within the same region. For instance, the prevalence of Chikungunya in Itang (Gambella) was approximately triple that of Lare (Gambella). This result is in line with previous studies that revealed the geographical region contributed to the inconsistency in Chikungunya prevalence (23). This is likely due to the extensive distribution and high population density of vectors, particularly in urban centers which are factors that favored their occurrence. The prevalence was higher in the town, especially in internally displaced populations sites, where various water containers such as tyres, clay pots, barrels, plastic water tanks, flower vases, and old cars are widely present as potential breeding grounds for Aedes mosquitoes (24). Additionally, the remoteness of certain areas may also play a role, as there are inequalities in the distribution of health facilities. These findings underscore the importance of considering the geographical context when implementing control and prevention strategies.

The current systematic review and meta-analysis demonstrated considerable variability in seroprevalence of Chikungunya among occupations in Ethiopia. The highest seroprevalence of Chikungunya was found among farmers compared to individuals in other occupations (Endale et al., 2020). This finding is consistent with a study conducted in northwest Ethiopia by Ferede et al. (2021) (see Figure7). The variation may be associated with the degree of exposure to vectors transmitting CHIKV. Seropositivity for Chikungunya is higher in individuals who regularly move in forests, engage in agricultural activities, and have documented incidents of mosquito bites (25). The fact that arboviruses typically circulate in forested areas through a sylvatic cycle, involving primates as reservoir hosts (25). The study by Thiberville et al. (2013), who reported that most seropositive individuals were engaged in farming activities, supports the higher prevalence of Chikungunya among farmers.

The prevalence of CHIKV in the reports included in this systematic review and meta-analysis was higher in 31–40 years of age-group. Similar findings were reported from Tanzania (26). The higher infection rate in this age group may suggest that people in those age groups are active workers and exposed to the bites of the vector that transmit the diseases. However, one study by Geleta et al. (2019) reported a higher Chikungunya prevalence in the age group of 5-14 years and with the lowest prevalence in the age group of ≤5 years. This difference may probably be a result of the sample size difference that is the number of people aged 5-14 years who underwent diagnosis was almost doubled that of the people with ≥30 years.

In this systematic review and meta-analysis, Education was associated with seroprevalence of Chikungunya in Ethiopia. People who were attended formal education were highly affected than those did not attended formal education. This may be associated with educated people living in the city or town where favorable environment for the vectors are found such as containers with water. Educated people might have frequent migration history compared to non-educated people (6,25). Moreover, higher prevalences in educated individuals might be associated with better access to healthcare facilities as they are living in the city where better health infrastructures are present, leading to increased detection and reporting of Chikungunya cases.

The results between studies were inconsistent with gender. The prevalence of Chikungunya in males was higher compared to females according to Asebe et al. (2021) and Ferede et al. (2021) (Figure 8). These findings are in line with other studies that revealed that men are more susceptible than women (27,28). This may affirm the argument that males face a higher likelihood of encountering mosquito bites in the course of agricultural work or other comparable travel and occupational activities. In contrast, Endale et al. (2020) reported that females were more CHIKV IgG+ compared to males indicated that females were more susceptible than males. This opposite trend is supported by other seroprevalence studies (29,30), where females were more susceptible than males. These conflicting reports highlight the necessity for further exploration of the associations between arbovirus infection and gender.

The results of subgroup analysis based on districts and test types were inconsistent across studies. The subgroup analysis, categorized by the type of tests conducted, unveiled differences in study outcomes. Districts employing ELISA demonstrated significant variability among the studies compared to districts that utilized RT-qPCR which exhibited lack of variability among the studies, though not statistical significance. The overall heterogeneity was found to be high. These findings underscore the importance of accounting for the type of test in the analysis, as it appears to be a significant factor contributing to the observed heterogeneity. Further exploration into the sources of variability and careful consideration of the clinical implications are essential for a comprehensive interpretation of these results.

### Study Strength and Limitations

The strengths of this systematic review and meta-analysis are that the study included published data since the first detection of Chikungunya, and it is the first to report the pooled seroprevalence of Chikungunya in Ethiopia.

However, the study has several limitations. The number of included studies is limited due to a restricted pool of available research, and the pooled prevalence may not accurately reflect the current reported rates. Almost all the included studies were cross-sectional. No molecular studies were carried out at the country level, except for RT-qPCR tests for confirmation. This limitation makes it challenging to predict the circulating strains of Chikungunya virus.

### Conclusions and Recommendations

The pooled prevalence of Chikungunya reveals that emerging and re-emerging diseases remain persistent public health concerns in Ethiopia, a country with limited resources and health infrastructure. Discrepancies in study results may be attributed to variations in sample size, the type of test performed, location, education, gender, and age. Additionally, the participating communities may exhibit diverse habits, behaviors, occupations, traditions, and local practices that could either increase or decrease their likelihood of exposure to the virus.

These findings underscore the importance of considering different factors and types of tests used for virus detection. Recognizing the significance of proactive operational readiness in mitigating the outbreak and spread of infectious diseases is recommended. Furthermore, it is crucial for the Ministry of Health and other concerned bodies to emphasize collaboration and public awareness campaigns to better respond to similar outbreaks.

### Author contributions

**DGG**: conceptualization, formulation of the data accusation methods, analyzed the data, prepared the draft manuscript, review and editing. **ATG**: collect data from published literatures, review and editing. **MZK**: collect data from published literatures, review and editing. **ABB:** collect data from published literatures, review and editing. **HD:** collect data from published literatures, review and editing. **BD:** collect data from published literatures, review and editing. **SLA**: collect data from published literatures, analyzed the data, review and editing.

## Abbreviations

CHIKV: Chikungunya Virus
ELISA: Enzyme Linked Immunosorbent Assay
IgG: Immunoglobulin G
IgM: Immunoglobulin M
RT-qPCR: quantitative Reverse-transcriptase polymerase chain reaction
WHO: World Health Organization

## Acknowledgements

Not applicable.

## Data availability

All data generated or analyzed during this study are included in this document.

## Ethics approval and consent to participate

Not applicable.

